# Reliability and validity of the hidden hunger assessment scale for Chinese high school students

**DOI:** 10.1101/2022.11.17.22282438

**Authors:** Ning Zhang, Minao Wang, Yuchen Zhang, Huake Cao, Yang Yang, Yuhang Shi, Yang Pei, Feixiang Yang, Yinan Du

**Author notes:** **Correspondence:** Yinan Du. Ning Zhang, Minao Wang, and Yuchen Zhang contributed equally to this work.

## Abstract

**Background:** Hidden hunger, also known as insufficient micronutrient intake, is still a severe public health problem, affecting over 2 billion people worldwide. High school students are in an important stage of growth and development. Due to huge academic stress, high school students often lack nutrition knowledge and have unhealthy eating habits, suggesting a higher risk for hidden hunger. However, information on the hidden hunger risk of high school students is limited, and tools for assessing hidden hunger are inadequate.

**Objectives:** To revise the hidden hunger assessment scale for high school students (HHAS-HSS) in China and assess its reliability and validity.

**Methods:** Based on the literature review, expert consultation, pre-experiment, and formal survey, a hidden hunger assessment scale was revised for high school students. The formal survey involved 9,336 high school students in 11 of the 16 cities in Anhui Province, China, from 1 September 2020 to 31 December 2020, and 9,038 valid questionnaires were collected and included in the analysis. The item analysis, internal consistency reliability, test-retest reliability, content validity, exploratory factor analysis, and confirmatory factor analysis of the HHAS-HSS were examined.

**Results:** The HHAS-HSS included a total of 4 dimensions and 12 items: “vegetables and food diversity” (3 items), “fruits and dairy products” (3 items), “micronutrient-dense foods” (4 items), and “health condition and eating habits” (2 items). The results showed a Cronbach’s alpha of 0.758, a split-half reliability of 0.829, and a test-retest reliability of 0.793, indicating good internal consistency. The exploratory factor analysis presented a four-factor model of the HHAS-HSS (KMO=0.820, P<0.001). Using the maximum variance rotation method, 4 factors were obtained, and the cumulative variance explained rate was 57.974%. Confirmatory factor analysis also supported the division of the scale into four dimensions and the above results showed that the construct validity of the scale has reached an acceptable level.

**Conclusions:** The HHAS-HSS is an effective and convenient tool for the assessment of hidden hunger with good reliability and validity.

## 1 Introduction

Nutrition during adolescence has a great influence on their growth and development, setting foundations for individual’s health in later life[1]. Due to the rapid development of worldwide economy, the lifestyle and dietary structure of people have undergone dramatic changes. An increasing number of adolescents suffer from overweight, obesity, and its related comorbidities, and micronutrient deficiency is still present today[1,2]. Hidden hunger usually refers to individuals who meet their energy requirements for consumption but have insufficient micronutrient intake, which typically exhibits deficiencies in microelements such as zinc, iron, iodine, folic acid, and vitamin A[3-5]. It has been well-established that hidden hunger can be associated with a wide range of diseases, including obesity, diabetes, cancer, osteoporosis, anemia, and mental and physical fatigue[6-12]. Therefore, understanding the hidden hunger status of individuals has positive influences on health promotion and disease prevention. However, owing to the limitations of detection tools and inconspicuous symptoms, the onset of hidden hunger is insidious and usually overlooked.

Currently, hidden hunger remains a severe challenge public health problem. More than 2 billion people worldwide suffer from micronutrient deficiency, especially in low- and middle-income countries across Asia, Africa, and the Americas[3-5,13,14]. Another study estimated that over half of pre-school children and two-thirds of non-pregnant women were in the state of micronutrient deficiency worldwide[3]. In China, challenges of dietary imbalance and hidden hunger still persist[5,15]. Approximately 300 million people were in a state of hidden hunger, and the inadequate intake of micronutrients among urban and rural residents was still widespread[15-19]. More than 50% of adults had below-average intake of vitamin A, thiamine, and vitamin C, and most adults had insufficient riboflavin and calcium intake [15]. In addition, micronutrient inadequacy deserves more attention especially for adolescents, which not only because the growth and development of adolescents rely on adequate nutrition support but also because they are leaders of the future[20]. Adequate intake of micronutrients contributes to brain development, bone health, immune system strengthening, and disease prevention[21-25]. However, adolescents often lack nutrition knowledge, and most of them have unhealthy eating habits and unbalanced dietary patterns[26-28], and this phenomenon has become increasingly serious during the COVID-19 epidemic[4,29-32]. Therefore, most adolescents may have a substantial risk for hidden hunger.

Notably, although micronutrients are crucial for adolescents to remain healthy, this problem appears to have been largely ignored[33,34]. Only a few manuscripts have focused on the hidden hunger of adolescents, and the data are still limited[20,35]. Additionally, the assessment of micronutrient intake mainly relies on invasive samples and instruments, which are expensive and unsuitable for mass screening. There is a need for an efficient and convenient assessment scale for evaluating the hidden hunger risk of adolescents. To that end, the main objective of this study is to develop the hidden hunger assessment scale for high school students (HHAS-HSS) in China and assess its reliability and validity. It is hoped that this study can better understand the current situation of hidden hunger among high school students and provide a useful tool for assessing the nutritional status of high school students.

## 2 Materials and methods

### 2.1 Study Design and Participants

The study was developed in four phases. First, based on the original scale, we conducted a literature review to revise the original entries of the hidden hunger assessment scale for high school students. Second, expert consultation was conducted to identify the items and scale structure. Third, pre-experiment was carried out to preliminarily verify the reliability and validity of the HHAS-HSS. Fourth, we performed a formal survey and assessed the reliability and validity of the scale.

#### 2.1.1 Literature review

Search terms such as “hidden hunger”, “micronutrient deficiency”, and “lack of micronutrient” were used for literature review using PubMed (https://pubmed.ncbi.nlm.nih.gov/). Corresponding Chinese words of these terms were used for the Chinese database CNKI (https://www.cnki.net/). Then, we removed duplicate literature, read the title and abstract of the literature, extracted the entries and scales related to the assessment of hidden hunger, and added or deleted the entries to initially form the HHAS-HSS for high school students.

#### 2.1.2 Expert consultation

A total of six experts from Anhui Medical University were selected as consulting objects, and their working years were more than five years. Through e-mail, expert consultation questionnaires were sent to the experts, and the content included the introduction of the research content, the instructions for filling in the form, and the content of HHAS-HSS. Experts were asked to evaluate the plausibility and correctness of these items, and the score range was 1 to 4 points (1 = not relevant/not clear, 4 = very relevant/very clear). These scores were used to calculate the Content Validity Index (I-CVI), Scale-level Content Validity Index (S-CVI), unanimous S-CVI (S-CVI/UA), and average S-CVI (S-CVI/AVE). The last column of each item in the questionnaire was the suggestion column, which was convenient for experts to provide comments and suggestions. At the end of the questionnaire, there was a supplementary recommendation column that could be used for the addition of entries. It is generally believed that the I-CVI is not less than 0.78, S-CVI/UA is not less than 0.8, and S-CVI/AVE is not less than 0.9, which indicates that the content is valid for application.

#### 2.1.3 Pre-experiment

The pre-experiment surveyed a total of 170 students in a high school in a city of Anhui Province. The characteristics of the participants were presented in **Supplementary table 1**. To check the comprehensibility of the HHAS-HSS, all the questionnaires were administered in face-to-face interviews. The data of pre-experiment were used to initially evaluate the performance of the questionnaires. The results indicated a Cronbach’s alpha coefficient of 0.755 and a split-half reliability coefficient of 0.826.

Using the extreme grouping method, there were significant differences between the high-level and low-level subdivisions for all the items (P<0.05). For exploratory factor analysis, the result (KMO = 0.776, P<0.001) indicated good appropriateness. All the factor loadings of the items were above 0.3. Except for the items in the “health condition and eating habits” dimension, all the item-total correlation coefficients ranged from 0.427 to 0.745 (P<0.01). For the items “Suffer from diarrhea, constipation and/or chronic gastric diseases” (an item-total correlation coefficient of 0.233) and “High study pressure, fast and irregular eating habits” (an item-total correlation coefficient of 0.227), we still decided to keep these entries considering the importance of micronutrient absorption for evaluation of hidden hunger. In addition, all item-dimension correlation coefficients ranged from 0.613 to 0.874 (P<0.01). The results suggested that all the items of the revised scale were reasonably designed and that none of the items were deleted.

#### 2.1.4 Formal survey

The formal survey was conducted from 1 September 2020 to 31 December 2020 in Anhui Province, China. Among 16 cities, 11 cities were sampled from Anhui Province according to their geographical locations and economic status, including Huangshan, Xuancheng, Wuhu, Hefei, Chuzhou, Anqing, Huainan, Huaibei, Suzhou, Fuyang, and Bengbu. Using the multistage stratified cluster random sampling method, 3 or 4 schools were randomly selected from each of the 11 selected cities. A total of 9,336 students participated in this survey and 9,038 valid questionnaires were included in the analysis. To verify the test-retest reliability, 58 students were randomly selected and re-surveyed after two weeks. The questionnaire survey was conducted anonymously. Informed consent was obtained from all participants (guardian). Ethical clearance was provided by the ethics committee of Anhui Medical University.

### 2.2 Instrument

The HHAS-HSS was revised based on the hidden hunger assessment scale of “Caring for Physicians Nutrition and Health Project” jointly sponsored by the Chinese Medical Doctor Association, Chinese Center for Disease Control and Prevention, Physicians Online, and Global Times[36]. The original scale consists of 12 items. After the literature review and expert consultation, the item “purchase of vegetables more than 3 times a week” was deleted and the item “Consumption of over 150 g of fish, poultry, eggs, and lean meat per day” was added. In addition, some items were reworded or modified according to the experts’ suggestions. The pre-experiment preliminarily revealed the acceptable construct, validity, and reliability of the scale. The HHAS-HSS consists of four dimensions: “vegetable and food diversity”, “fruit and dairy product”, “micronutrient-dense foods”, and “health condition and eating habits”. Each item of the scale has three options: “often”, “sometimes”, and “rarely”, assigning a score of 5, 3, 1, or 1, 3, 5. The evaluation criteria are as follows: total score ≥ 50 points, “good condition, no shortage”; 49 to 39 points, “may lack, need to improve”; and total score ≤ 38 points, “lack”. The Chinese version and English version of HHAS-HSS are presented in **Supplementary table 2**.

### 2.3 Data collection

The design of this study was approved by the school authorities, and the questionnaires were distributed through our trained investigators. All the participants were taught how to fill in the scales and encouraged to answer truthfully. After the quality audit, unqualified and incomplete questionnaires were eliminated. A double-entry pattern was adopted by two researchers and the data were cross-checked to ensure correct entry using Epidata 3.1 software (EpiData Association, Odense, Denmark).

### 2.4 Statistical analysis

Data analysis was performed using SPSS 25.0 (IBM, Armonk, NY) and AMOS 26.0 (IBM, Armonk, NY). The demographic characteristics of participants were described using frequencies and percentages. For item analysis, the extreme grouping method, item-dimension correlations, and item-total correlations were performed. The correlation coefficients exceeding 0.3 were acceptable. Cronbach’s alpha coefficients, split-half reliability, and test-retest reliability (odd-even method) were used for examining the reliability. Reliability values exceeding 0.7 were considered acceptable. For validity analysis, I-CVI, S-CVI, S-CVI/UA, and S-CVI/AVE were calculated to estimate content validity. Exploratory factor analysis using the maximum variance orthogonal rotation method and confirmatory factor analysis were conducted to examine construct validity. The criteria of fit indices from confirmatory factor analysis were set as follows: χ²/df<8.00, GFI>0.90, AGFI>0.80, PGFI>0.50, NFI>0.90, IFI>0.90, TLI>0.90, CFI>0.90, and RMSEA<0.08.

## 3 Results

### 3.1 Demographic characteristics

A total of 9,336 high school students participated in this survey. After quality control, 9,038 valid questionnaires were collected and included in the analysis, and the effective rate was 96.8%. Among the 9,038 subjects, there were 4,537 (50.2%) male students and 4,501 (49.8%) female students, with 4,071 (45.0%) participants from senior high school level one, 3,296 (36.5%) from senior high school level two, and 1,671 (18.5%) from senior high school level three. Of all the students, 2,577 (28.5%) came from urban areas and 6,461 (71.5%) came from rural areas. In addition, 2,773 (30.7%) participants were from the only child family, and the other 6,225 (69.3%) participants came from families with more than one child. The demographic characteristics of the subjects are presented in **Table 1**.

**Table 1.**
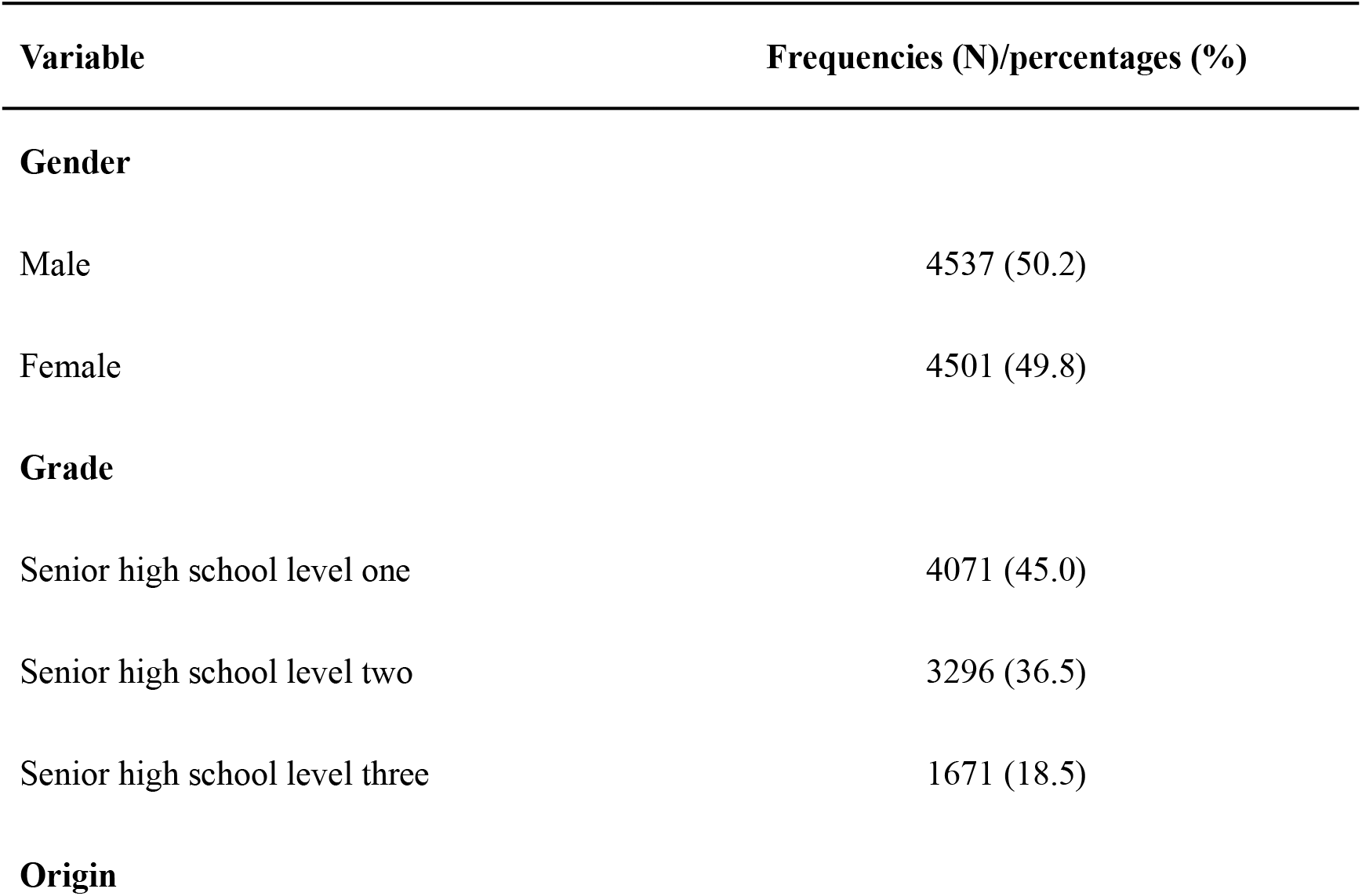
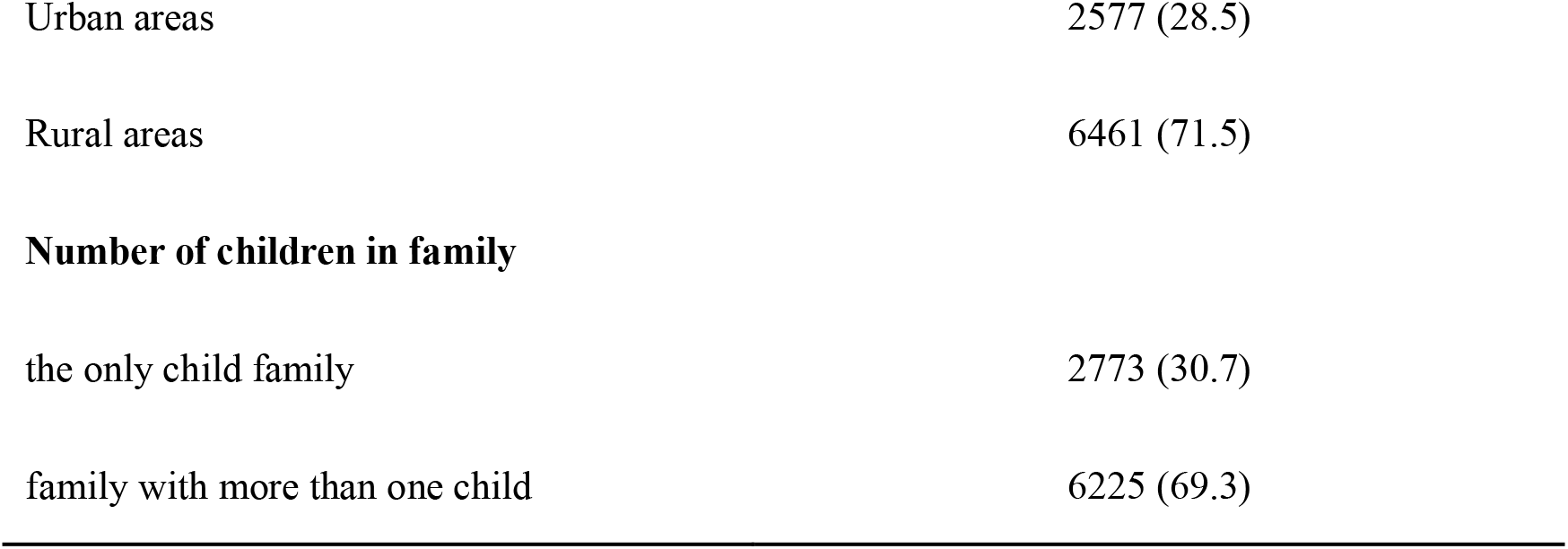
Demographic characteristics of the subjects (N=9,038)

### 3.2 Item analysis

The item discrimination of the scale was analyzed using the extreme grouping method, with 27% as the threshold. The questionnaires were arranged in descending order according to the total score. The top 2,440 scales were set as the high-level group, and the bottom 2,440 were set as the low-level group. The results of the independent samples t-test suggested that scores of all the items were significantly different between the high-level and low-level subdivisions (P<0.05). Each item was also tested using item-dimension correlations and item-total correlations. The results revealed that all item scores were significantly correlated with the total score of both the scale and the corresponding dimension (P<0.01). Except for one item, the item-total correlation coefficients ranged from 0.310 to 0.699, and item-dimension correlation coefficients ranged from 0.653 to 0.858, indicating a good correlation (**Table 2**).

**Table 2.** Item-total correlations and item-dimension correlations (N=9,038)

| Items | item-dimension | item-total | Items | item-dimension | item-total |
| --- | --- | --- | --- | --- | --- |
|  | correlations | correlations |  | correlations | correlations |
| A1 | 0.676** | 0.505** | C1 | 0.678** | 0.553** |
| A2 | 0.831** | 0.619** | C2 | 0.666** | 0.527** |
| A3 | 0.794** | 0.612** | C3 | 0.653** | 0.560** |
| B1 | 0.855** | 0.692** | C4 | 0.673** | 0.448** |
| B2 | 0.858** | 0.699** | D1 | 0.769** | 0.210** |
| B3 | 0.702** | 0.540** | D2 | 0.819** | 0.310** |

\*\* p<0.01

### 3.3 Reliability Analysis

The results showed a Cronbach’s alpha coefficient of 0.758 for the HHAS-HSS and the split-half reliability coefficient was 0.829. The calculation of the internal consistency reliability was repeated when one item was deleted, and the Cronbach’s alpha coefficient if item deleted remained fairly stable ranging from 0.717-0.777. After two weeks, a total of 58 students were randomly selected and asked to completed the scales. The test-retest reliability was calculated as 0.793. These results suggested good internal consistency and reliable reliability of the HHAS-HSS.

### 3.4 Validity Analysis

#### 3.4.1 Content Validity

Six experts were invited to evaluate the revised scale. The results showed I-CVI indices ranging from 0.833 to 1.000, an index of 0.986 for S-CVI/AVE, and an index of 0.917 for S-CVI/UA, indicating a good content validity.

#### 3.4.2 Exploratory Factor Analysis

The KMO was computed to be 0.820, the chi-square value of Bartlett’s spherical test was 22683.3, and the degree of freedom was 66, with P<0.001. Based on these results, an exploratory factor analysis is appropriate to perform. The maximum variance method was used to obtain four rotated factors to explain 57.974% of the total variance, and the factor loadings ranged from 0.428 to 0.830 (all >0.4). These results showed stable structural validity. The details of the exploratory factor analysis are shown in **Table 3**.

**Table 3.** Exploratory factor analysis of HHAS-HSS (N=9,038)

| Items | Factors |  |  |  |
| --- | --- | --- | --- | --- |
|  | Factor 1 | Factor 2 | Factor 3 | Factor 4 |
| A1 | 0.603 |  |  |  |
| A2 | 0.795 |  |  |  |
| A3 | 0.731 |  |  |  |
| B1 |  | 0.830 |  |  |
| B2 |  | 0.827 |  |  |
| B3 |  | 0.586 |  |  |
| C1 |  |  | 0.662 |  |
| C2 |  |  | 0.545 |  |
| C3 |  |  | 0.428 |  |
| C4 |  |  | 0.711 |  |
| D1 |  |  |  | 0.796 |
| D2 |  |  |  | 0.759 |
| Cumulative variance contribution rate (%) | 11.102 | 41.357 | 49.913 | 57.974 |

#### 3.4.3 Confirmatory factor analysis

For confirmatory factor analysis, four-factor models were evaluated using different indices (**Figure 1**). The results of the fitting of the structural equations were χ²=1417.656, χ²/df=29.534, GFI=0.974, AGFI=0.958, PGFI=0.600, NFI=0.938, IFI=0.940, TLI=0.917, CFI=0.939, and RMSEA=0.056 (**Table 4**). Because the value of χ²/df is proportioned with the sample size, it is unsuitable as a reference index in our study. Except for χ²/df, all other parameters were located within the ideal interval, indicating that the model had a good fit.

**Table 4.** The results of model fitting (N=9,038)

| Fitting index | Fitting standard | Modified result |
| --- | --- | --- |
| $\chi^2$ | the smaller the better | 1417.656 |
| $\chi^2/\text{df}$ | <8.00 | 29.534 |
| GFI | >0.90 | 0.974 |
| AGFI | >0.80 | 0.958 |
| PGFI | >0.50 | 0.600 |
| NFI | >0.90 | 0.938 |
| IFI | >0.90 | 0.940 |
| TLI | >0.90 | 0.917 |
| CFI | >0.90 | 0.939 |
| RMSEA | <0.08 | 0.056 |

**Figure 1.**
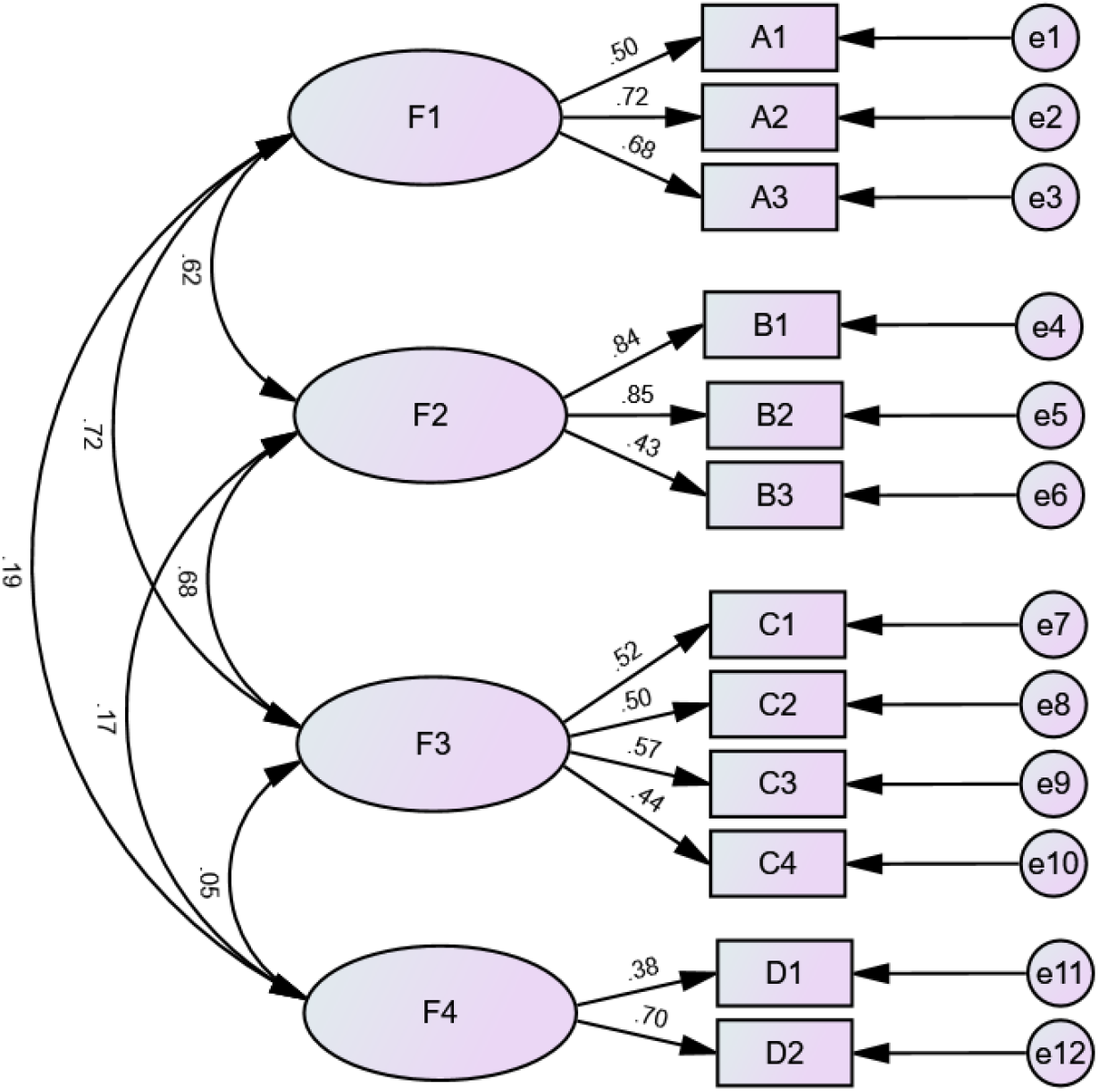
Confirmatory factor analysis of the four-factor model of the HHAS-HSS (N=9,038).

## 4. Discussion

Currently, methods of combating hidden hunger included optimized food system strategies, biofortification, food-to-food fortification, knowledge intervention, micronutrient replacement, and nutritional supplements[37-42]. However, although positive policies have been introduced and a decreasing trend has appeared, hidden hunger is still a major public health problem across the world[3,13,43-45]. Because of the inconspicuous symptoms, the prevalence of hidden hunger worldwide might be underestimated. Existing surveillance methods for hidden hunger usually include invasive procedures with a need for blood samples, which have the disadvantages of high cost, puncture trauma, and complex procedures, limiting large-scale screening of hidden hunger[46,47]. In this study, we developed a hidden hunger assessment scale for high school students named HHAS-HSS, providing a convenient, effective, and reliable tool for investigating micronutrient deficiency.

The HHAS-HSS consists of four dimensions and 12 items. Except for one item, the results of item-total correlations and item-dimension correlations were in the range of 0.310 to 0.858, reflecting good correlations. In addition, using the extreme grouping method, the scores of the high-level and low-level groups presented significant differences, indicating a good degree of differentiation. The results of the reliability analyses showed a Cronbach’s alpha coefficient of 0.758 and a split-half reliability coefficient of 0.829. The test-retest reliability was calculated as 0.793. The above results indicated that the scale had good reliability. To verify the content validity, six experts from Anhui Medical University were invited as consulting objects. The results showed that the I-CVI was in the range of 0.833-1.000, the S-CVI/AVE was 0.986, and the S-CVI/UA was 0.917, reflecting good content validity. For the structural validity of the scale, the results of the exploratory factor analysis showed a KMO value of 0.820 (P<0.001). The factor rotation was performed using the maximum variance orthogonal rotation method, and the scale was divided into four factors, with a cumulative variance interpretation rate of 57.974%. The factor loadings of each entry were in the range of 0.428-0.830. Confirmatory factor analysis supported the conclusion and the model fit indices were acceptable. The above results showed that the scale had good structural validity.

To our knowledge, we are the first to report a hidden hunger assessment scale especially for high school students. Previous studies have revealed a positive correlation between dietary diversity and micronutrient adequacy, and dietary diversity indicators including dietary diversity scores (DDS) and food insecurity (FI) assessment tools including household food security survey (HFSS) have been used for screening hidden hunger[48-50]. However, the possible impact of bioaccessibility also needs to be considered in the assessment, which seems to be neglected in previous research[42]. The bioaccessibility of a compound is its ability to be absorbed from the gastrointestinal tract after it has been released from its matrix[51]. In our scale, we developed the dimension “health condition and eating habits”, which included two items: “Suffer from diarrhea, constipation and/or chronic gastric diseases” and “High study pressure, fast and irregular eating habits”. The unhealthy state of digestive system, eating disorders, and unhealthy eating habits reduce digestion and absorption of nutrients and lead to micronutrient deficiency and malnutrition[52-57]. In summary, the results indicate that the HHAS-HSS presents good reliability and validity among high school students in Anhui Province, China, and is a new and effective tool for assessing the risk of hidden hunger.

However, this study also has some limitations. The study was conducted among high school students in Anhui Province, China. The representativeness of participants is limited and studies with larger sample sizes are needed for further confirmation. In addition, due to the differences in economic level and regional eating habits between different regions, countries, and races, the universal applicability of the scale needs to be explored. Finally, some participants might have performed error assessment due to memory bias.

Overall, the HHAS-HSS is a valid and reliable instrument to evaluate the risk of hidden hunger among high school students. Compared with past screening methods, the scale has the characteristics of low cost, convenient operation, time savings and labor savings, without the need for invasive procedures. Our study provides a powerful tool for the public health or education sectors to monitor the prevalence of hidden hunger and evaluate interventions.

## Data Availability

The original contributions presented in the study are included in the article/Supplementary Material, further inquiries can be directed to the corresponding author.

## 6 Author Contributions

Conceptualization: N.Z.; data curation: N.Z. and H.C.; formal analysis: M.W., Y.Z., H.C., and Y.Y.; funding acquisition: N.Z. and Y.D., investigation: N.Z., M.W., Y.Z., H.C., Y.Y., Y.S., Y.P., and F.Y.; methodology: N.Z., M.W., and Y.D.; project administration: N.Z., H.C., and Y.D.; resources: N.Z. and Y.D.; software: N.Z., M.W., and Y.Z.; supervision: N.Z. and Y.D.; validation: N.Z., M.W., Y.Z., H.C., Y.Y., Y.S., Y.P., F.Y., and Y.D.; visualization: N.Z., M.W., and Y.Z.; writing – original draft: N.Z., M.W., and Y.Z.; writing – review & editing: N.Z. and Y.D.

## 8 Acknowledgments

The authors thank all the participants for their contribution to the research. We also thank the Mathematical Medicine Integration Innovation Training Program for Undergraduate Students (MITUS) for valuable guidance, support, and providing research opportunities and resources.

## 10 Ethics Statement

Approval was obtained from the ethics committee of Anhui Medical University. All the participants (guardian) expressed informed consent. The procedures used in this study adhere to the tenets of the Declaration of Helsinki.

